# Improved diagnosis of COVID-19 vaccine-associated myocarditis with cardiac scarring identified by cardiac magnetic resonance imaging

**DOI:** 10.1101/2024.03.20.24304640

**Authors:** Josephine Warren, Daryl Cheng, Nigel W Crawford, Bryn Jones, Rui Lun Ng, Annette Alafaci, Dion Stub, Philip Lew, Andrew Taylor

**Affiliations:** Department of Cardiology, Alfred Hospital, Melbourne, Victoria, Australia; Central Clinical School, Monash University, Melbourne, Victoria, Australia; Department of General Medicine, Royal Children’s Hospital, Melbourne, Victoria, Australia; SAEFVIC, Infection and Immunity and Global Health, Murdoch Children’s Research Institute, Melbourne, Victoria, Australia; Centre for Health Analytics, Melbourne Children’s Campus, Melbourne, Victoria, Australia; Department of Paediatrics, Faculty of Medicine, Dentistry and Health Sciences, University of Melbourne, Melbourne, Victoria, Australia; Heart Research Group, Murdoch Children’s Research Institute, Melbourne, Victoria, Australia; Department of Cardiology, Royal Children’s Hospital, Melbourne, Victoria, Australia; School of Public Health and Preventive Medicine, Monash University, Victoria, Australia; Department of Radiology, Alfred Hospital, Melbourne, Victoria, Australia

**Author notes:** **Funding:** Nil. **Disclosures:** Nil. **Address for Correspondence:** Professor Andrew J. Taylor, The Alfred Hospital, 55 Commercial Road, Melbourne, Victoria, Australia 3004.

## Abstract

**Background:** Myocarditis is a rare but potentially serious complication of COVID-19 vaccination. Cardiac magnetic resonance (CMR) late gadolinium enhancement (LGE) imaging can identify cardiac scar, which may improve diagnostic accuracy and prognostication.

**Objectives:** To define the incidence of long-term LGE post COVID-19 vaccine-associated myocarditis (C-VAM) and to establish the additive role of CMR in the diagnostic work-up.

**Methods:** Patients with Brighton Collaboration Criteria Level 1 (definite) or Level 2 (probable) C-VAM were prospectively recruited from the Surveillance of Adverse Events Following Vaccination In the Community (SAEFVIC) database to undergo CMR at least 12 months after diagnosis. As there were limited patients with access to baseline CMR, prior CMR results were not included in the initial case definition. The presence of LGE on follow-up CMR was then integrated into the diagnostic algorithm and the reclassification rate (definite vs. probable) was calculated.

**Results:** Sixty-seven patients with C-VAM (mean age 30 ± 13 years, 72% male) underwent CMR evaluation. Median time from vaccination to CMR was 548 (range 398-603) days. Twenty patients (30%) had persistent LGE, most frequently found in the basal inferolateral segment (n = 11). At diagnosis, nine patients (13%) were classified as definite and 58 (87%) as probable myocarditis. With integration of CMR LGE data, 16 patients (28%) were reclassified from probable to definite myocarditis.

**Conclusion:** Persistent LGE on CMR occurs in one third of patients with C-VAM. Without CMR at diagnosis, almost one third of patients are misclassified as probable rather than definite myocarditis.

## Introduction

The rapid and timely development of effective vaccines against COVID-19 resulted in the prevention of an estimated 14 million deaths worldwide within the first 12 months of their introduction(1). As of January 2024, over 69 million doses of COVID-19 vaccine have been administered in Australia(2).

Myocarditis is a rare but important complication of COVID-19 vaccination, with an incidence of approximately 2 cases per 100,000(2). Although the clinical course of COVID-19 vaccine-associated myocarditis is usually benign and self-limiting(3, 4), previous studies evaluating the use of cardiac magnetic resonance imaging (CMR) in the acute setting have found imaging abnormalities are common. These findings include late gadolinium enhancement (LGE), which reflects myocardial fibrosis and scarring (seen in up to 90% of cases) and elevated myocardial T2 relaxation time, which is indicative of myocardial oedema (in up to 80% of cases)(5, 6).

The Brighton Collaboration Case Definition(7) for the diagnosis of Level 1 (definite) myocarditis requires either histological evidence of myocarditis, or the combination of elevated myocardial biomarkers in association with diagnostic imaging features on either CMR or transthoracic echocardiography (TTE). Endocardial biopsies were rarely undertaken in Australia to confirm a C-VAM diagnosis, and due to barriers with funding and access, most patients did not undergo evaluation with CMR at the time of diagnosis, which meant diagnostic classification and therefore risk stratification depended primarily on clinical and echocardiographic characteristics alone. This may have resulted in both diagnostic uncertainty and the potential failure to identify those at higher long-term risk of cardiac complications.

Whilst little is known about the longer-term sequelae of this condition, specifically the impact on cardiac function and its clinical correlation, it is particularly crucial to define as COVID-19 vaccine-associated myocarditis is predominantly a disease of the young and healthy(8). Current evidence is limited to only a few small cohort studies with a relatively short follow up period.

Thus, we present our study evaluating long-term CMR findings in a cohort of 67 adolescents and adults with COVID-19 vaccine-associated myocarditis, which is, to our knowledge, the largest such study with the longest follow up period. In addition, we sought to highlight the critical role of CMR in the work up of this condition by defining the reclassification rate of probable to definite myocarditis with the addition of CMR with LGE imaging to baseline clinical and echocardiographic assessment.

## Methods

### Patient selection

All participants were prospectively recruited from a centralised surveillance database containing patients who received a diagnosis of COVID-19 vaccine-associated myocarditis between August 2021 and March 2022.

All adverse vaccine related events in Victoria, Australia are reported to SAEFVIC (Surveillance of Adverse Events Following Vaccination In the Community), the state-wide vaccine safety service. SAEFVIC has been operating since 2007 and comprises central reporting enhanced passive and active surveillance systems for all vaccine adverse events following immunisation. All identified reports of myocarditis or myopericarditis were systematically followed up to obtain clinical information to allow independent categorisation of COVID-19 vaccine-associated myocarditis according to the Brighton Criteria (see below)(7). Based on available information, all myocarditis cases were categorised as Level 1 (definite) or Level 2 (probable) and were included in this study.

Patients were contacted via telephone and invited to participate in the study. Patients who had never had a CMR or had an initial abnormal CMR demonstrating LGE, inflammation or left ventricular dysfunction were invited to undergo follow up CMR at either the Alfred Hospital or the Royal Children’s Hospital in Melbourne, Australia. All patients provided written consent prior to participation. An additional subset of patients who had a normal CMR following diagnosis were included in the study but did not undergo a follow up CMR. Exclusion criteria prohibiting enrolment for follow up CMR included severe renal impairment (estimated glomerular filtration rate <30ml/min/1.73m^2^), allergy to gadolinium contrast, pregnancy, current breastfeeding and any other contraindication to magnetic resonance imaging (MRI).

All procedures were performed with approval from the Alfred Hospital Human Research Ethics Committee (HREC 484/22) and Research Ethics and Governance at the Royal Children’s Hospital (HREC 80233).

### Brighton Criteria definitions(7)

Prior to the COVID-19 pandemic, there was no universally accepted vaccine safety surveillance case definition for myocarditis (or pericarditis) as an adverse event following immunisation. The Brighton Collaboration established a consensus case definition for vaccine-related myocarditis with distinct levels of diagnostic certainty, which was published in 2021.

According to the Brighton Criteria, the diagnosis of COVID-19 vaccine-associated myocarditis requires symptom onset within two weeks of COVID-19 vaccination.

A diagnosis of Level 1 (definite) myocarditis requires either histological evidence of myocarditis on endomyocardial biopsy, or the combination of elevated myocardial biomarkers (troponin I or T) and diagnostic imaging features on either CMR or TTE(7). Diagnostic CMR findings include the presence of either patchy myocardial oedema on T2 mapping or LGE on T1 weighted images involving ≥1 segments in a non-coronary distribution. Diagnostic TTE findings include new right or left ventricular dysfunction (either global or segmental), new regional wall motion abnormalities, new left ventricular diastolic dysfunction, ventricular dilatation, or change in ventricular wall thickness.

Level 2 (probable) myocarditis is defined as the combination of cardiac symptoms (including chest pain, palpitations, dyspnoea, diaphoresis or sudden death) and either elevated cardiac biomarkers (troponin I or T, or CK-myocardial band), diagnostic TTE findings or new abnormalities on ECG (including ST-segment or T-wave abnormalities, evidence of atrial or ventricular arrhythmia, or conduction disease)(7).

For our initial case definition, we divided our cohort into baseline Brighton Criteria Level 1 or 2 on the basis of clinical and echocardiographic data alone. We believed this best reflected the current clinical practice given access to CMR at the time of diagnosis was limited in Australia. We then included follow up CMR data into reclassification, specifically the presence of LGE, which can be assumed in the absence of prior cardiac events to represent scarring from prior vaccine myocarditis. This enabled an evaluation of the additive role of CMR in the diagnosis and classification of COVID-19 vaccine-associated myocarditis.

### Alfred Hospital Cardiac Magnetic Resonance Imaging protocol

All prospective adult CMR scans were performed at the Alfred Hospital on the same MRI scanner (Siemens Magnetom Skyra, Germany) according to a standardised CMR imaging protocol.

Patients were scanned supine with ECG gating. Following the acquisition of three planar scout views (axial, coronal and sagittal), standard 4-, 3- and 2-chamber long-axis views were obtained. Subsequently, one pre-contrast short-axis T1 mapping slice at the mid-ventricle level and one T2 mapping slice at the same level were acquired.

IV gadolinium was then administered. Left ventricular (LV) function was assessed by a standard steady state free precession technique (SSFP) using a short axis stack covering the whole ventricle followed by a T1 optimisation sequence.

LGE imaging was performed 10-15 minutes following contrast injection using an inversion recovery gradient echo technique on the same short axis stack utilised for cine imaging. 4-, 3- and 2-chamber long axis views were taken to evaluate the presence and location of LGE.

One post-contrast short-axis T_1_ mapping slice at the mid-ventricular level was acquired, as well as a 3-slice short axis short tau inversion recovery (STIR) slices.

### Royal Children’s Hospital Cardiac Magnetic Resonance Imaging protocol

All paediatric CMR scans were performed at the Royal Children’s Hospital on a 1.5T Siemens Magnetom Aera scanner (Siemens Medical Solutions, Erlangen, Germany). A standardised myocarditis protocol was used.

Patients were scanned in the supine position with ECG gating. Following acquisition of localiser images (axial, coronal and sagittal) a SSFP stack was obtained for anatomical assessment. Subsequently SSFP 4-chamber and 2-chamber cine images were obtained. Three pre-contrast T1 mapping image slices were obtained at the left ventricular base, mid-ventricle and apex. A stack of T2 mapping images was obtained from base to apex.

IV gadolinium was administered at this point. A short axis stack of SSFP cine images was obtained from base to apex for assessment of ventricular function.

LGE imaging was performed 8-12 minutes post gadolinium administration using a free-breathing, motion-corrected phase-sensitive inversion recovery sequence. 4-chamber and short axis images (base to apex) were obtained. Three post-contrast T1 mapping image slices were obtained at the left ventricular base, mid-ventricle and apex.

### Cardiac Magnetic Resonance Imaging analysis

All follow-up adult CMRs were performed at the same tertiary cardiac centre and were read by two expert CMR specialists.

All follow-up paediatric CMRs were performed at the same tertiary paediatric cardiac centre and were analysed by two independent CMR specialists who arrived at a consensus conclusion for all studies.

Patients with normal baseline imaging were included in the cohort but did not undergo follow up CMR.

### Statistical analysis

Continuous variables are expressed as mean ± standard deviation (SD) or median (interquartile range), and categorical variables are expressed as number (percentage).

Categorical variables were compared using Pearson’s chi-square test.

Statistical analysis was performed using Stata/MP 14.2 for Windows (College Station, TX, USA). P values <0.05 were considered to represent statistical significance.

## Results

A total of 67 patients underwent CMR evaluation following a diagnosis of COVID-19 vaccine-related myocarditis.

The baseline and treatment characteristics are summarised in Table 1. The mean age was 30 ± 13 years and the majority of patients were male (72%). Most patients (55 of 67 patients, 82%) developed myocarditis following the Comirnaty® BNT162b2 COVID-19 (Pfizer-BioNTech) vaccine. Ten patients (15%) had the Spikevax® mRNA-1273 (Moderna) vaccine prior to symptoms, while two patients received a diagnosis of myocarditis following the Vaxzevria ChAdOx1-S (AstraZeneca) vaccine (3%). The second COVID-19 vaccine dose was the most common precipitant for myocarditis in this cohort, in 47 patients (70%). Most patients received a single type of vaccine (n = 63, 94%), while a small proportion (n = 4, 6%) received a combination of types (all four cases received two doses of AstraZeneca, followed by a dose of Pfizer).

**Table 1.**
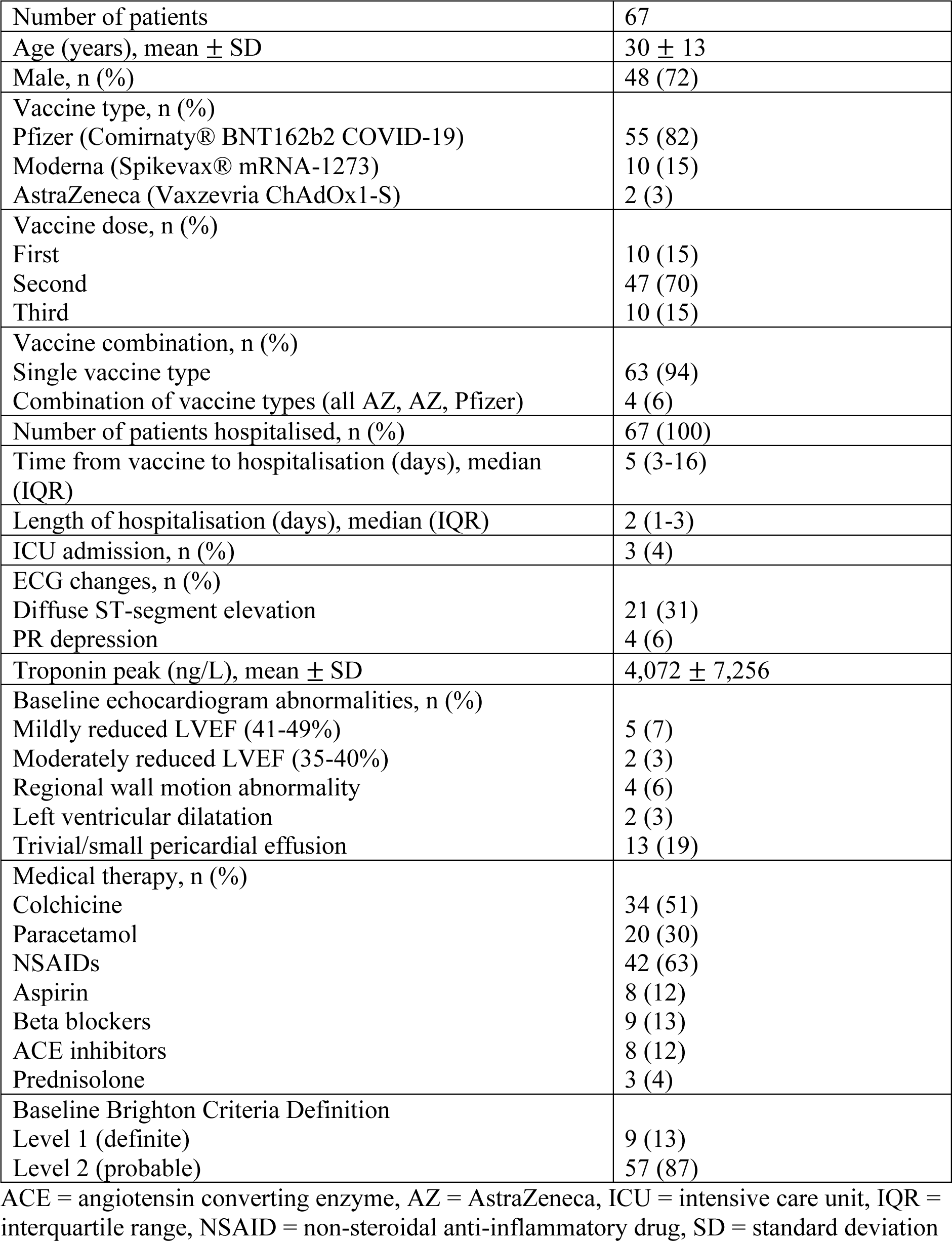
Baseline and Treatment Characteristics.

|  |  |
| --- | --- |
| Number of patients | 67 |
| Age (years), mean ± SD | 30 ± 13 |
| Male, n (%) | 48 (72) |
| Vaccine type, n (%) |  |
| Pfizer (Comirnaty® BNT162b2 COVID-19) | 55 (82) |
| Moderna (Spikevax® mRNA-1273) | 10 (15) |
| AstraZeneca (Vaxzevria ChAdOx1-S) | 2 (3) |
| Vaccine dose, n (%) |  |
| First | 10 (15) |
| Second | 47 (70) |
| Third | 10 (15) |
| Vaccine combination, n (%) |  |
| Single vaccine type | 63 (94) |
| Combination of vaccine types (all AZ, AZ, Pfizer) | 4 (6) |
| Number of patients hospitalised, n (%) | 67 (100) |
| Time from vaccine to hospitalisation (days), median (IQR) | 5 (3-16) |
| Length of hospitalisation (days), median (IQR) | 2 (1-3) |
| ICU admission, n (%) | 3 (4) |
| ECG changes, n (%) |  |
| Diffuse ST-segment elevation | 21 (31) |
| PR depression | 4 (6) |
| Troponin peak (ng/L), mean ± SD | 4,072 ± 7,256 |
| Baseline echocardiogram abnormalities, n (%) |  |
| Mildly reduced LVEF (41-49%) | 5 (7) |
| Moderately reduced LVEF (35-40%) | 2 (3) |
| Regional wall motion abnormality | 4 (6) |
| Left ventricular dilatation | 2 (3) |
| Trivial/small pericardial effusion | 13 (19) |
| Medical therapy, n (%) |  |
| Colchicine | 34 (51) |
| Paracetamol | 20 (30) |
| NSAIDs | 42 (63) |
| Aspirin | 8 (12) |
| Beta blockers | 9 (13) |
| ACE inhibitors | 8 (12) |
| Prednisolone | 3 (4) |
| Baseline Brighton Criteria Definition |  |
| Level 1 (definite) | 9 (13) |
| Level 2 (probable) | 57 (87) |

ACE = angiotensin converting enzyme, AZ = AstraZeneca, ICU = intensive care unit, IQR = interquartile range, NSAID = non-steroidal anti-inflammatory drug, SD = standard deviation

All 67 patients were hospitalised. The median time from vaccination to hospitalisation was five days (interquartile range (IQR) 3-16 days). The length of hospitalisation was usually brief, with a median duration of two days (IQR 1-3 days). Only three patients (4%) required intensive care admission. Abnormal ECG findings were seen in 29 (43%) patients at presentation, with the most common abnormality being diffuse ST-segment elevation, which was present in 21 patients. The mean peak troponin was 4072 ± 7256 µg/L. A total of 10 (15%) patients had abnormal echocardiographic changes at presentation, seven of whom had reduced left ventricular ejection fraction (LVEF <50%). Four patients had regional wall motion abnormalities (two without associated reduced LVEF) and two patients had left ventricular dilation (one without other associated abnormalities on TTE). A further 13 patients (19%) had a trivial or small pericardial effusion.

Approximately half the patients received colchicine as initial therapy (51%), with 42 patients (63%) treated with non-steroidal anti-inflammatory agents. Only three patients (4%) received corticosteroids. No patients required inotropic support, mechanical ventilation, or extra-corporeal membrane oxygenation. On baseline assessment (without CMR), nine patients (13%) were classified as definite myocarditis, while the remaining 58 patients (87%) were classified as probable myocarditis.

Table 2 contains the CMR results. Of the 67 patients had CMR, 60 studies were performed at >12 months post vaccination. A total of 44 prospective CMR scans were performed on adults who either had no initial CMR or an abnormal baseline CMR. A further 16 prospective CMR scans were performed on adolescents (aged <18) for the same reason. Seven patients had a CMR performed as part of their initial workup that was normal, so in these patients follow up CMR was not performed. The median time from vaccination to follow up CMR was 548 days (IQR 398-603).

**Table 2.** Cardiac Magnetic Resonance Imaging Results.

|  |  |
| --- | --- |
| Number of patients with CMR | 67 |
| No. patients with follow up CMR (>12 months), n (%) | 60 (90) |
| Time from vaccine to follow up CMR (days), median (IQR) | 548 (398-603) |
| **Findings** |  |
| Normal CMR, n (%) | 47 (70) |
| Myocardial late gadolinium enhancement (LGE), n (%) | 20 (30) |
| Pattern of LGE, n |  |
| Midwall | 7 |
| Subendocardial | 2 |
| Subepicardial | 13 |
| Elevated STIR (> 2.2 adults), n % | 0 (0) |
| STIR, mean ± SD | 1.67 ± 0.29 |
| Abnormal T1 time (>1050ms adults, >1067ms children), n (%) | 0 (0) |
| T1 time (ms), mean ± SD | 992 ± 89 |
| Elevated T2 time (> 60ms adults, >50ms children), n (%) | 0 (0) |
| T2 time (ms), mean ± SD | 48 ± 6 |
| Left ventricular ejection fraction |  |
| Normal (>55%) | 59 (88) |
| Low-normal (50-54%) | 8 (12) |
| Mildly reduced (41-49%) | 0 |
| Moderately reduced (35-40%) | 0 |
| Severely reduced (<35%) | 0 |
| Left ventricular regional wall motion abnormality (RWMA) | 2 (3) |
| Anteroapical RWMA | 1 (50) |
| Mid inferior and inferolateral RWMA | 1 (50) |
| Pericardial effusion, n (%) | 8 (12) |
| Trivial/small | 8 (100) |
| Moderate + | 0 |
| Baseline Level 1 (definite) |  |
| LGE | 4 (44%) |
| No LGE | 5 (56%) |
| Baseline Level 2 (probable) |  |
| LGE | 16 (28%) |
| No LGE | 41 (72%) |

CMR = Cardiac Magnetic Resonance Imaging, IQR = interquartile range, LGE = Late Gadolinium Enhancement, LV = left ventricular, RWMA = regional wall motion abnormality, STIR = short-tau inversion recovery

Almost a third of patients (n = 20, 30%) had evidence of persistent myocardial late gadolinium enhancement, and the common territory containing LGE was the basal inferolateral wall (n = 11) (Figure 1). The pattern of LGE was most commonly subepicardial (n = 13), followed by midwall (n = 7) and then subendocardial (n = 2). Two patients had left ventricular regional wall motion abnormalities which corresponded to the territory of LGE. Eight patients had low-normal ejection fraction (LVEF 50-54%) on follow up CMR, two of whom had mild or moderately reduced LVEF on baseline echocardiogram. The remaining five patients with reduced LVEF on TTE had normalisation of their ejection fraction on follow up CMR. All patients had a normal T1 and T2 time on follow up CMR. The mean T1 time was 992 ± 89ms and the mean T2 time was 48 ± 6ms. A total of eight patients (12%) had a pericardial effusion, all of which were trivial or small and of no haemodynamic significance.

**Figure 1.**
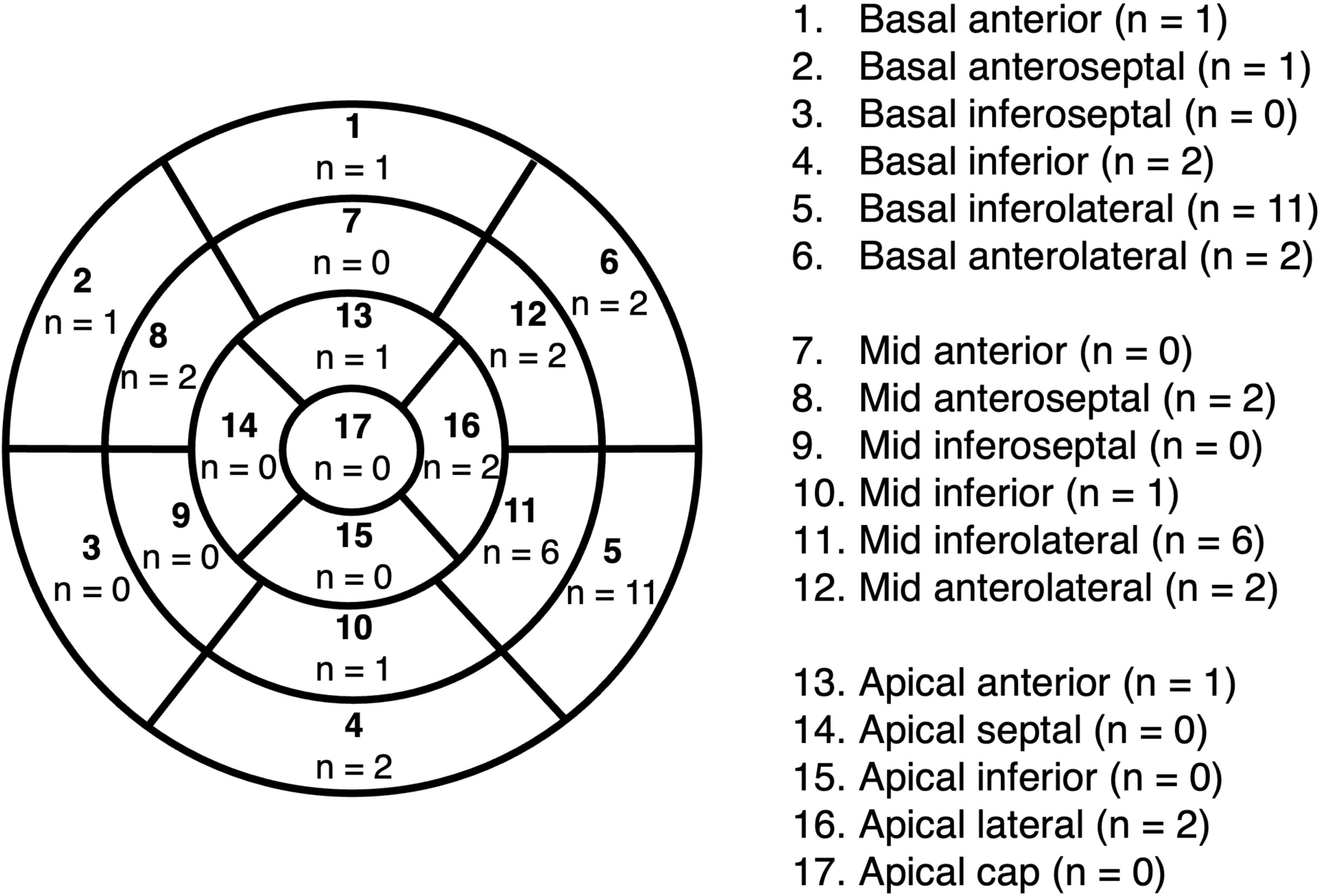
Distribution of left ventricular late gadolinium enhancement. Distribution of late gadolinium enhancement according to the 17-segment model of the left ventricle. The most common location for late gadolinium enhancement on follow-up CMR was the basal inferolateral segment. CMR = cardiac magnetic resonance imaging

Four patients (44%) who were originally classified as definite myocarditis had LGE on follow up CMR. Sixteen patients (28%) who were originally classified as probable myocarditis had LGE, which resulted in re-classification to definite myocarditis (Figure 2). There was no significant difference in the rate of LGE between those who were originally classified as definite or probable myocarditis (44% vs. 28%, p = 0.30).

**Figure 2.**
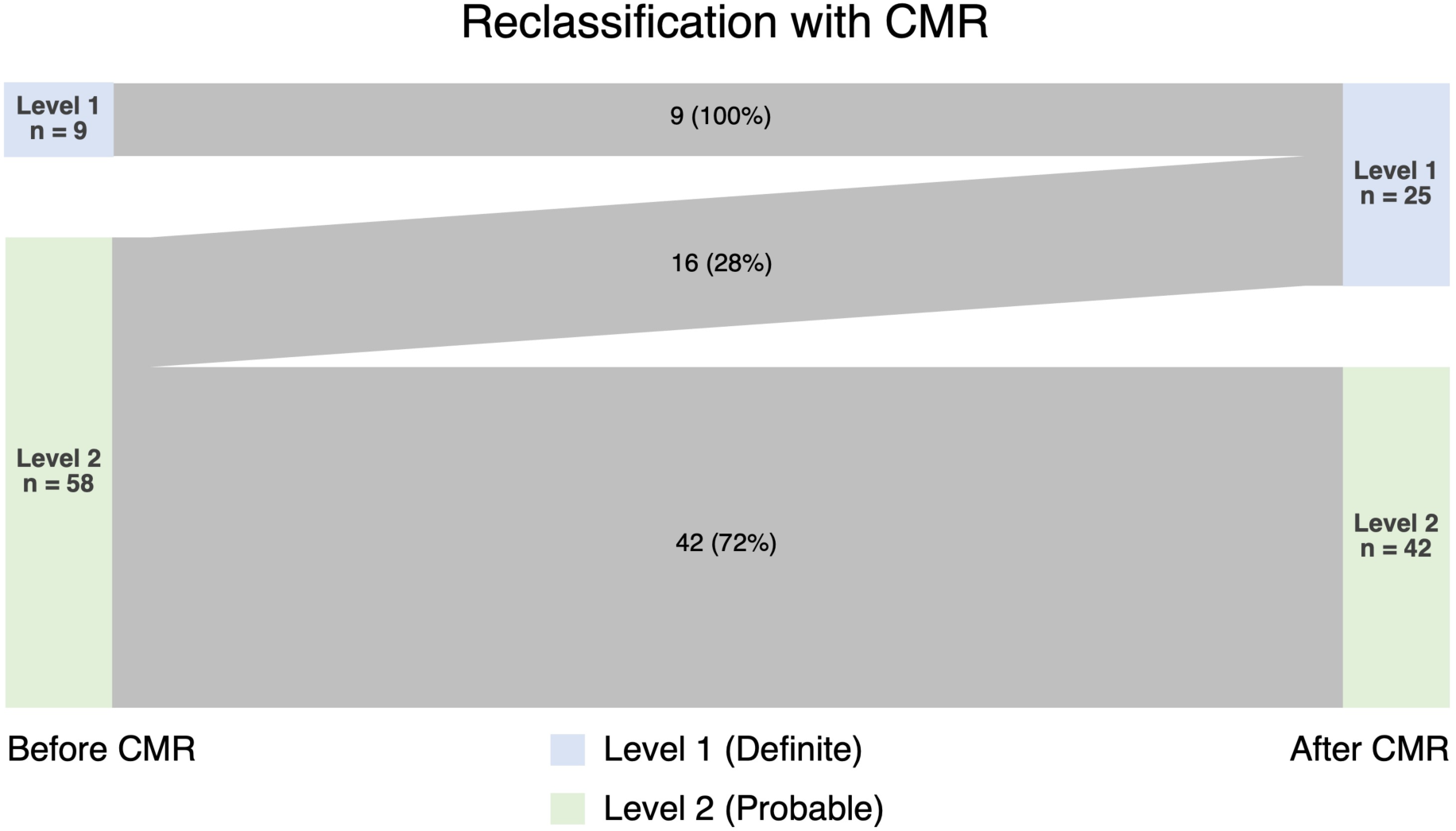
Sankey Diagram demonstrating Brighton Criteria reclassification based on late gadolinium enhancement on follow-up CMR. Almost a third of patients (16 of 56, 28%) who were originally classified as probable myocarditis were found to have persistent late gadolinium enhancement on CMR, resulting in reclassification to definite myocarditis. CMR = cardiac magnetic resonance imaging

Table 3 demonstrates the results of baseline and follow up CMR for the 20 patients who underwent serial studies. The median time from vaccination to baseline CMR was 48 days (range 14-143) and the median time from baseline to follow up CMR was 457 days (range 400-556). On baseline CMR, 19 (95%) patients had late gadolinium enhancement. Eight patients (20%) had evidence of myocardial oedema identified by hyperintensity on T2-weighted imaging. Two patients had an elevated T1 time. Five patients had low-normal ejection fraction (LVEF 50-54%). Of the 19 patients with LGE at baseline, 10 patients had resolution of the LGE on follow up CMR, five patients had persistent LGE but to a lesser extent and four patients had unchanged LGE on follow up CMR. No patients had progression of LGE on follow up CMR.

**Table 3.** Comparison of Baseline to Follow up CMR.

|  |  |
| --- | --- |
| Number of patients with baseline and follow up CMR | 20 |
| Time from vaccine to baseline CMR (days), median (IQR) | 48 (14-143) |
| Time between baseline and follow up CMR (days), median (IQR) | 457 (400-556) |
| Baseline CMR findings, n (%) |  |
| Late gadolinium enhancement | 19 (95) |
| Elevated T1 time | 2 (10) |
| Hyperenhancement on T2 weighted images | 8 (40) |
| Normal left ventricular ejection fraction | 15 (75) |
| Low-normal left ventricular ejection fraction | 5 (25) |
| Left ventricular regional wall motion abnormalities | 0 |
| Comparison to LGE on follow up CMR, n (%) |  |
| No longer visible | 10 (53) |
| Reduced in size | 5 (26) |
| Unchanged | 4 (21) |
| Increased in size | 0 |

CMR = Cardiac Magnetic Resonance Imaging, IQR = interquartile range, LGE = Late Gadolinium Enhancement

## Discussion

In this study we describe the long-term CMR findings in a cohort of patients with COVID-19 vaccine-associated myocarditis. This is the largest study of its kind to date with the longest follow up period, with a median time from vaccination to CMR of over 18 months. We found that the incidence of persistent myocardial fibrosis is high, seen in almost a third of patients at >12 months post diagnosis, which could have implications for the management and prognosis of this predominantly young cohort. Furthermore, our findings highlight the critical role played by CMR in the diagnosis and risk-stratification of this condition. Without CMR, almost a third of patients were misclassified as probable rather than definite myocarditis.

Consistent with previous studies, our study population was comprised of mostly younger male patients in whom the clinical course of myocarditis was relatively mild. Nonetheless, incidence of LGE was high, seen in 30% of the total cohort. The long-term clinical implications of LGE in this condition are as yet unknown, but LGE has been demonstrated to confer worse prognosis in non-COVID-19 vaccine-associated myocarditis(9, 10), especially if it persists beyond six months(11). Furthermore, a study of 222 patients with biopsy-proven myocarditis identified that LGE on CMR was the strongest predictor of long-term mortality (hazard ratio 8.4 for all-cause mortality, hazard ratio 12.8 for cardiac mortality), and carried more prognostic significance than LVEF, degree of LV dilation and New York Heart Association functional status(12). This prognostic impact persists at 10-year follow up(13). Concerningly, left ventricular fibrosis is also a nidus for ventricular arrhythmia in chronic cardiomyopathies(14–16). As such, the identification of LGE on CMR in patients with a history of COVID-19 vaccine-associated myocarditis should at a minimum prompt regular clinical review, to allow for periodic monitoring for arrhythmias and deterioration in cardiac function.

A recent study of 12-month follow-up CMR in 40 adolescents with C-VAM found a comparable incidence of persistent LGE to our study, with a rate of 37.5%(17) (seen in 15 out of 40 patients). Researchers similarly found a predilection of LGE for the lateral LV segments. We describe a lower incidence of LGE than a previous study of 16 adolescents with a history of C-VAM who underwent serial CMR evaluation at diagnosis and between three and eight months post(6). Investigators found persistent LGE in 68% of patients, although they observed a significant reduction in the burden of LGE in all subjects. In this prior study, the follow-up CMR scans were performed at a very early time interval post-diagnosis, and it is possible that there may have been lower incidence of LGE if the CMR studies were performed >12 months.

Almost a third of patients (28%) who were originally diagnosed with probable myocarditis were found to have persistent LGE on CMR, resulting in reclassification to definite myocarditis. As such, clinical evaluation without CMR may lead to underestimation of long-term risk through failure to identify potentially prognostically significant LGE.

We would assert that on the basis of our findings, echocardiographic evaluation alone in a resource-available setting is insufficient in the diagnostic work up of COVID-19 vaccine-associated myocarditis. Characteristic echocardiographic findings required for a diagnosis of definite myocarditis (e.g. reduced LVEF or regional wall motion abnormalities) were rare in our cohort despite relatively high rates of persistent myocardial scarring, further emphasising the limitations of echocardiography in the evaluation of this condition. CMR has been well described as crucial to the diagnostic work up of patients with suspected non-COVID-19 vaccine-associated myocarditis(18, 19), exhibiting better diagnostic performance than all other modalities of assessment, including endomyocardial biopsy(20). In an Australian context, the widespread lack of access to and appropriate reimbursement for CMR at initial diagnosis potentially disadvantaged many patients with COVID-19 vaccine-associated myocarditis.

It must be noted that any discussion surrounding adverse events following immunisation requires acknowledgement of the overwhelmingly positive impact of the widespread global vaccination policy against COVID-19, which prevented an estimated 14 million deaths within the first 12 months of distribution(1). In this same period, a large study of >42 million vaccinated people in the United Kingdom found that the risk of myocarditis associated with COVID-19 infection significantly outweighed the risk associated with COVID-19 vaccination(21), a finding which was echoed in a subsequent meta-analysis by Voleti et al(22). Furthermore, a recent study by Lai et al indicates a favourable medium-term prognosis of C-VAM compared to viral myocarditis. Only one death was observed in a cohort of 104 patients with C-VAM, which conferred a 92% lower mortality risk than viral myocarditis(4). Of note, however, this study did not include CMR data, which prevented exploration of CMR predictors of adverse outcome (such as LGE).

Although this is the largest cohort study to date evaluating follow-up CMR in patients with COVID-19 vaccine-associated myocarditis, our study is nonetheless limited by its small sample size. Additionally, it does not contain clinical follow up to ascertain correlation between CMR findings and symptoms or adverse events. For this reason, even longer-term follow up studies are required to define the prognostic significance of LGE in this condition. We did not collect clinical data on the development of interval cardiac events between vaccination and follow up CMR, therefore it is technically possible that the LGE observed could have been related to a different cardiac condition. Finally, participation in our study was voluntary, which may have resulted in selection bias and impacted the accuracy of the overall incidence of long-term LGE.

In conclusion, long-term myocardial fibrosis is a common finding in patients with COVID-19 vaccine-associated myocarditis. CMR enables accurate diagnostic assessment and risk stratification of patients with this condition and is critical to a comprehensive work up and long-term follow up strategy.

## Data Availability

Data available on reasonable request.

## Abbreviations

CMR: Cardiac Magnetic Resonance Imaging
COVID-19: Coronavirus disease of 2019
C-VAM: COVID-19 vaccine-associated myocarditis
LGE: Late gadolinium enhancement
MRI: Magnetic Resonance Imaging
TTE: Transthoracic echocardiography
SAEFVIC: Surveillance of Adverse Events Following Vaccination In the Community

## Acknowledgements

Dr Josephine Warren is supported by National Health and Medical Research Council (Australia) and Monash University Scholarships (Melbourne, Australia). Professor Dion Stub is supported by National Health and Medical Research Council and National Heart Foundation Fellowships (Australia). Professor Andrew Taylor is supported by a National Health and Medical Research Council Investigator Fellowship (Australia).

## Conflicts of Interest

Nothing to Disclose.

## Notes

### Competing Interest Statement

The authors have declared no competing interest.

### Funding Statement

This study did not receive any funding.

### Author Declarations

The ethics committees of the Royal Children's Hospital and Alfred Hospital name gave ethical approval for this work.

## References

1. Watson OJ, Barnsley G, Toor J, Hogan AB, Winskill P, and Ghani AC, Global impact of the first year of COVID-19 vaccination: a mathematical modelling study. The Lancet Infectious Diseases, 2022. 22(9): 1293–302.

2. Australian Government Department of Health and Aged Care, COVID-19 Vaccine Rollout. 2024.

3. Witberg G, Barda N, Hoss S, et al., Myocarditis after Covid-19 Vaccination in a Large Health Care Organization. New England Journal of Medicine, 2021. 385(23): 2132–9.

4. Lai FTT, Chan EWW, Huang L, et al., Prognosis of Myocarditis Developing After mRNA COVID-19 Vaccination Compared With Viral Myocarditis. Journal of the American College of Cardiology, 2022. 80(24): 2255–65.

5. Fronza M, Thavendiranathan P, Chan V, et al., Myocardial Injury Pattern at MRI in COVID-19 Vaccine–associated Myocarditis. Radiology. 0(0): 212559.

6. Schauer J, Buddhe S, Gulhane A, et al., Persistent Cardiac Magnetic Resonance Imaging Findings in a Cohort of Adolescents with Post-Coronavirus Disease 2019 mRNA Vaccine Myopericarditis. The Journal of Pediatrics. 254: 233–7.

7. Sexson Tejtel SK, Munoz FM, Al-Ammouri I, et al., Myocarditis and pericarditis: Case definition and guidelines for data collection, analysis, and presentation of immunization safety data. Vaccine, 2022. 40(10): 1499–511.

8. Therapeutic Goods Administration, COVID-19 vaccine weekly safety report -26-09-2023. Australian Government Department of Health and Aged Care., 2023.

9. Yang F, Wang J, Li W, et al., The prognostic value of late gadolinium enhancement in myocarditis and clinically suspected myocarditis: systematic review and meta-analysis. European Radiology, 2020. 30(5): 2616–26.

10. Georgiopoulos G, Figliozzi S, Sanguineti F, et al., Prognostic Impact of Late Gadolinium Enhancement by Cardiovascular Magnetic Resonance in Myocarditis. Circulation: Cardiovascular Imaging, 2021. 14(1): e011492.

11. Aquaro GD, Ghebru Habtemicael Y, Camastra G, et al., Prognostic Value of Repeating Cardiac Magnetic Resonance in Patients With Acute Myocarditis. J Am Coll Cardiol, 2019. 74(20): 2439–48.

12. Grün S, Schumm J, Greulich S, et al., Long-term follow-up of biopsy-proven viral myocarditis: predictors of mortality and incomplete recovery. J Am Coll Cardiol, 2012. 59(18): 1604–15.

13. Greulich S, Seitz A, Müller KAL, et al., Predictors of Mortality in Patients With Biopsy-Proven Viral Myocarditis: 10-Year Outcome Data. Journal of the American Heart Association, 2020. 9(16): e015351.

14. Gutman SJ, Costello BT, Papapostolou S, et al., Reduction in mortality from implantable cardioverter-defibrillators in non-ischaemic cardiomyopathy patients is dependent on the presence of left ventricular scar. European Heart Journal, 2018. 40(6): 542–50.

15. Disertori M, Rigoni M, Pace N, et al., Myocardial Fibrosis Assessment by LGE Is a Powerful Predictor of Ventricular Tachyarrhythmias in Ischemic and Nonischemic LV Dysfunction: A Meta-Analysis. JACC: Cardiovascular Imaging, 2016. 9(9): 1046–55.

16. Kuruvilla S, Adenaw N, Katwal AB, Lipinski MJ, Kramer CM, and Salerno M, Late gadolinium enhancement on cardiac magnetic resonance predicts adverse cardiovascular outcomes in nonischemic cardiomyopathy: a systematic review and meta-analysis. Circ Cardiovasc Imaging, 2014. 7(2): 250–8.

17. Yu CK, Tsao S, Ng CW, et al., Cardiovascular Assessment up to One Year After COVID-19 Vaccine-Associated Myocarditis. Circulation, 2023. 148(5): 436–9.

18. Petersen SE, Friedrich MG, Leiner T, et al., Cardiovascular Magnetic Resonance for Patients With COVID-19. JACC Cardiovasc Imaging, 2022. 15(4): 685–99.

19. Ferreira VM, Plein S, Wong TC, et al., Cardiovascular magnetic resonance for evaluation of cardiac involvement in COVID-19: recommendations by the Society for Cardiovascular Magnetic Resonance. J Cardiovasc Magn Reson, 2023. 25(1): 21.

20. Eichhorn C, Greulich S, Bucciarelli-Ducci C, Sznitman R, Kwong RY, and Gräni C, Multiparametric Cardiovascular Magnetic Resonance Approach in Diagnosing, Monitoring, and Prognostication of Myocarditis. JACC Cardiovasc Imaging, 2022. 15(7): 1325–38.

21. Patone M, Mei XW, Handunnetthi L, et al., Risk of Myocarditis After Sequential Doses of COVID-19 Vaccine and SARS-CoV-2 Infection by Age and Sex. Circulation, 2022. 146(10): 743–54.

22. Voleti N, Reddy SP, and Ssentongo P, Myocarditis in SARS-CoV-2 infection vs. COVID-19 vaccination: A systematic review and meta-analysis. Front Cardiovasc Med, 2022. 9: 951314.

